# Proteogenomic and observational evidence implicate ANGPTL4 as a potential therapeutic target for colorectal cancer prevention

**DOI:** 10.1101/2024.11.20.24317649

**Authors:** J. Yarmolinsky, M.A. Lee, E. Lau, F. Moratalla-Navarro, E.E. Vincent, R. Li-Gao, P.C.N. Rensen, K.W. van Dijk, K.K. Tsilidis, A. Sangphukieo, E. Ebrahimi, J. Hampe, L. Le Marchand, F.J.B. van Duijnhoven, K. Visvanathan, M.O. Woods, M. Guevara, S. Sieri, G. Masala, K. Papier, S. Virani, T. Dudding, A. Dehghan, D. Wang, V. Moreno, M.J. Gunter, I. Tzoulaki

## Abstract

**Background:** Preclinical, observational, and genetic epidemiological evidence implicate circulating lipids in cancer development. The role of approved and emerging lipid-perturbing medications in cancer risk is unclear.

**Patients and methods:** We employed *cis*-Mendelian randomization (MR) and colocalisation to evaluate the role of 5 lipid-perturbing drug targets (ANGPTL3, ANGPTL4, APOC3, CETP, PCSK9) in risk of 5 cancers (breast, colorectal, head and neck, ovarian, prostate) in up to 319,661 cases and 348,078 controls. We further triangulated findings using direct measures of pre-diagnostic protein targets in case-cohort analyses in the European Prospective Investigation into cancer and Nutrition (EPIC). To gain mechanistic insight into the role of ANGPTL4 in carcinogenesis, we examined the impact of the ANGPTL4 p.E40K loss-of-function variant on differential gene expression in normal colon tissue in the BarcUVa-Seq project. Finally, we evaluated the association of *ANGPTL4* gene expression in colon tumour tissue with all-cause mortality in The Cancer Genome Atlas (TCGA).

**Results:** In analysis of 78,473 cases and 107,143 controls, genetically-proxied circulating ANGPTL4 inhibition was associated with a reduced risk of colorectal cancer (OR per SD decrease: 0.76, 95% CI 0.66-0.89, *P* = 5.52 x 10^-4^, colocalisation posterior probability = 0.83). This association was replicated in the EPIC cohort using pre-diagnostic circulating ANGPTL4 concentrations in 977 incident colorectal cancer cases and 4,080 sub-cohort members (HR per log10 decrease: 0.92, 95% CI 0.85-0.99, *P* = 0.02). In gene set enrichment analysis of differential gene expression in 445 normal colon tissue samples, ANGPTL4 loss-of-function was associated with down-regulation of several biological pathways implicated in cancer (FDR *P* < 0.05), including those involved in cellular proliferation, epithelial-to-mesenchymal transition, and bile acid metabolism. In analysis of 465 colon cancer patients, lower *ANGPTL4* expression in tumour tissue was associated with reduced risk of all-cause mortality (HR per log2 decrease: 0.85, 95% CI 0.73-0.99; *P* = 0.04). There was little evidence of association of genetically-proxied inhibition of ANGPTL4 or other lipid targets with the other cancer outcomes evaluated.

**Conclusion:** Our integrative proteogenomic and observational analyses suggest a protective role of lower circulating ANGPTL4 concentrations in colorectal cancer risk. These findings support further evaluation of ANGPTL4, an emerging drug target for hypertriglyceridemia, as a potential therapeutic target for colorectal cancer prevention.

**Highlights:**

- We used complementary proteogenomic and observational analyses to investigate the effect of lipid-perturbing drug targets on cancer risk
- Across all methods there was consistent evidence for a protective role of lower circulating ANGPTL4 concentrations in colorectal cancer risk
- Our findings highlight the possibility of repurposing pharmacological ANGPTL4 inhibition as a novel approach for colorectal cancer prevention

## Introduction

Lipids perform various essential physiological functions including providing an energy reserve, serving as structural components of cell membranes, and participating in cellular signaling^1^. It is well established that select lipid parameters (e.g. LDL cholesterol) contribute to atherosclerotic cardiovascular disease (ASCVD). Along with their role in ASCVD, preclinical studies have suggested that lipids may also influence carcinogenesis through several mechanisms including those related to insulin resistance, inflammation, and oxidative stress^2^. Reprogramming of lipid metabolism also plays a critical role in promoting tumorigenesis and is considered an emerging hallmark of cancer^3,4^. For example, cancer cells must harness lipid metabolism to support cell division, adapt to stress, and enable metastatic dissemination^5–7^. In addition, lipid metabolic reprogramming can remodel the tumor microenvironment by influencing the recruitment, survival, and function of immune cells^8^.

Consistent with a role of circulating lipids in cancer development, preclinical and epidemiological studies have suggested that several lipid-perturbing medications may lower cancer risk^9–13^. For example, knockdown of ANGPTL3, the target of the lipid-lowering therapy evinacumab, has been shown to suppress proliferation, migration, and invasion in several cancer cell lines^14–16^. In addition, PCSK9 inhibition using siRNA, gene knockout, or anti-PCSK9 vaccination promotes apoptosis in *in vitro* cancer models^13^. Observational epidemiological studies have also reported that long-term statin users have lower rates of site-specific cancer as compared to non-users^12,17–21^.

These findings collectively suggest the potential for repurposing approved and/or emerging lipid-perturbing CVD medications for cancer prevention. However, in the absence of randomised clinical trial data, the causal nature of these medications in cancer onset, and thus their suitability as intervention targets, is unclear. This is because of the uncertain relevance of preclinical disease models to humans and the susceptibility of conventional observational analyses to residual confounding and reverse causation, undermining confident causal inference^22–24^.

Here, we leveraged four complementary epidemiological approaches to triangulate evidence on the potential causal role of lipid-perturbing drug targets in cancer risk. We employed drug-target Mendelian randomization (MR) to systematically evaluate the effect of 5 approved or emerging lipid-perturbing drug targets for CVD (APOC3, ANGPTL3, ANGPTL4, CETP, PCSK9) on risk of 5 cancers (breast, colorectal, head and neck, ovarian, prostate). This approach leverages the natural randomisation of germline genetic variants at meiosis and can minimise conventional epidemiological issues of confounding and reverse causation. We then examined the association of pre-diagnostic direct measures of circulating protein targets and cancer risk in case-cohort analyses in the European Prospective Investigation into Cancer and Nutrition (EPIC) study. To gain mechanistic insight into the role of ANGPTL4 in carcinogenesis, we explored the impact of ANGPTL4 loss-of-function on differential gene expression in normal colon tissue samples in the University of Barcelona and the University of Virginia Genotyping and RNA Sequencing (BarcUVa-Seq) project. Finally, to explore whether ANGPTL4 is involved in cancer progression, we evaluated the association of *ANGPTL4* gene expression in colon tumour tissue with all-cause mortality in The Cancer Genome Atlas (TCGA).

## Methods

A step-by-step overview of the analytical stages of this work is presented in **Figure 1**.

### Study populations

We selected cancer outcomes where there was prior evidence from Mendelian randomization studies suggesting a role of circulating lipids (i.e. breast, colorectal) or lipid-perturbing drug targets (i.e. head and neck, ovarian, prostate) in their aetiology^25–31^.

For drug-target MR analyses, summary genetic association estimates for overall and estrogen receptor (ER)-stratified breast cancer risk in up to 122,977 cases and 105,974 controls were obtained from the Breast Cancer Association Consortium (BCAC)^32^. Summary genetic association estimates for overall and site-specific colorectal cancer risk in up to 78,473 cases and 107,143 controls were obtained from an analysis of the Genetics and Epidemiology of Colorectal Cancer Consortium (GECCO), ColoRectal Transdisciplinary Study (CORECT), and Colon Cancer Family Registry (CCFR)^33^. Summary genetic association estimates for prostate cancer risk in up to 79,148 cases and 61,106 controls were obtained from the Prostate Cancer Association Group to Investigate Cancer Associated Alterations in the Genome (PRACTICAL) consortium^34^. Summary genetic association data for overall and subtype-specific head and neck cancer in up to 13,554 cases and 32,914 controls were obtained from a prior GWAS^35^. Summary genetic association data on overall and histotype-specific epithelial ovarian cancer risk were obtained from 25,509 cases and 40,941 controls from the Ovarian Cancer Association Consortium (OCAC)^36^.

For genetic instrument validation analyses, summary genetic association data on circulating LDL cholesterol, HDL cholesterol, and triglycerides were obtained from analyses of ∼1.3 million participants in the Global Lipids Genetics Consortium (GLGC) analysis^37^.

These analyses were restricted to participants of European ancestry. Further information on statistical analysis, imputation, and quality control measures for these studies is available in the original publications. All studies contributing data to these analyses had the relevant institutional review board approval from each country, in accordance with the Declaration of Helsinki, and all participants provided informed consent.

For triangulation analyses using direct measures of protein drug targets, we used data from the EPIC cohort study^38^. EPIC includes over 520,000 individuals who were recruited between 1992 and 2000 from 23 study centres across 10 European countries. Participants were 35-70 years of age at recruitment and approximately 70% of the cohort are women. The study design has been described previously^39^. We limited the present analyses to 10,261 individuals recruited into a multi-endpoint case-cohort within EPIC, of whom 6,876 had incident cancer, including 977 colorectal cancer cases (658 colon cancer, 319 rectal cancer). Further information on the case-cohort design employed is presented in **Supplementary Materials**. Incident first primary cancer cases were identified through a combination of centre-specific methods including health insurance records, cancer and pathology registries, and active follow-up through study participants and their next of kin. Follow-up for all individuals and events of interest began at recruitment and ended upon the occurrence of the event, loss to follow-up, or the last date of ascertainment, whichever came first. In the present study, cancer endpoints were defined as the first incident cancer diagnosis, using the following ICD-0-3 codes: colon: C180, C181, C182, C183, C184, C185, C186, C187, C188, C189; rectal (including rectosigmoid junction): C199, C209.

For analyses examining the association of tumour gene expression with all-cause mortality, we obtained gene expression (RNA-Seq), demographic, and clinicopathological data from 465 colon adenocarcinoma (TCGA-COAD) cases in TCGA. TCGA is a publicly available resource that has sequenced and molecularly characterised over 20,000 primary cancer and matched normal samples across 33 cancer types. Additional study details of TCGA have been described elsewhere^40^.

### Genetic instrument construction

Genetic instruments for circulating APOC3, ANGPTL3, ANGPTL4, and PCSK9 concentrations were constructed from genome-wide significant (*P* < 5 x 10^-8^) and independent (LD r^2^ < 0.001) single-nucleotide polymorphisms (SNPs) in or within 1MB from the gene encoding the relevant protein using summary genetic association data from a prior GWAS in 35,559 individuals of Icelandic ancestry^41^. Replication analyses were performed in an independent GWAS of 54,219 participants of primarily white British ancestry in the UK Biobank. For SNPs that replicated (*P* < 0.05) and were directionally consistent, SNP weights were obtained from UK Biobank analyses.

Such an approach mimics a “three-sample” MR design and has been shown to minimise bias from “Winner’s curse” in the presence of overlap of sample participants across “replication” and “outcome” data sources (i.e. UK Biobank participants in this analysis)^42^. Circulating APOC3 measures were not available in the UK Biobank and therefore no replication analyses were performed for this target. A genetic instrument for circulating CETP concentrations was constructed using genome-wide significant (*P* < 5 x 10^-8^) and independent (LD r^2^ < 0.001) SNPs associated with circulating CETP concentrations in or within 500KB from *CETP* in a GWAS of 5,706 participants in the Netherlands Epidemiology of Obesity (NEO) study^43^. In analyses exploring pathways mediating the effect of ANGPTL4, we also constructed a genetic instrument to proxy circulating triglyceride concentrations using genome-wide significant (*P* < 5 x 10^-8^) and independent (r^2^ < 0.001) SNPs associated with circulating triglycerides, irrespective of genomic position of variants, using data on ∼1.3 million participants in the previously described GLGC analysis.

### Drug-target Mendelian randomization primary and sensitivity analyses

For drug targets instrumented by a single SNP, the Wald ratio was used to generate effect estimates and the delta method was used to approximate standard errors. For drug targets instrumented by two or more SNPs, inverse-variance weighted (IVW) random-effects models (permitting overdispersion in models) were used to estimate causal effects^44^.

Drug-target MR can generate valid tests of the causal null hypothesis if the instrument used to proxy a drug target (i) is associated with the drug target (“relevance”); (ii) does not share a common cause with the outcome (“exchangeability”); and (iii) affects the outcome only through the drug target (“exclusion restriction”). Under the assumption of monotonicity, drug-target MR can provide valid point estimates for participants whose exposure is influenced by the instrument (i.e. a local average treatment effect).

We tested the “relevance” assumption by generating estimates of the proportion of variance of each drug target explained by the instrument (r^2^) and F-statistics. F-statistics can be used to evaluate if results are likely to be driven by weak instrument bias, i.e. reduced statistical power when an instrument explains a limited proportion of the variance in a drug target. As a convention, an F-statistic > 10 is used to indicate that instruments are unlikely to be vulnerable to weak instrument bias^45^.

We evaluated the “exclusion restriction” assumption by performing various sensitivity analyses. First, where applicable, we validated our instruments by evaluating the effect of genetically-proxied drug targets on downstream biomarkers influenced (i.e. for approved drugs) or presumed to be influenced (i.e. for emerging drugs) by the target as “positive control” analyses (i.e. triglyceride concentrations for APOC3, ANGPTL3, ANGPTL4; HDL cholesterol concentrations for CETP; LDL cholesterol concentrations for PCSK9)^25,46–49^. Second, colocalisation was performed to evaluate whether drug targets and both “positive control” lipid measures and cancer outcomes showing evidence of association in MR analyses (Bonferroni-corrected *P* < 6.17 x 10^-4^) were likely to share the same causal variant at a given locus. Such an analysis can permit evaluation of whether drug targets and disease endpoints are influenced by distinct causal variants that are in linkage disequilibrium (LD) with each other, indicative of horizontal pleiotropy (i.e. an instrument influencing an outcome through pathways independent to the exposure), a violation of the exclusion restriction assumption^50^. Colocalisation was performed by generating ± 100 kb windows around sentinel variants for drug targets using pair-wise conditional and colocalisation (PWCoCo) using default prior probabilities (*p1* = *p2* = 1 x 10^-4^, *p12* = 1 x 10^-5^). We used a posterior probability of colocalisation (PP_colocalisation_) > 0.80 to support colocalisation of drug targets and disease outcomes. Third, for analyses examining the association of circulating triglycerides with colorectal cancer risk, we employed three complementary “pleiotropy-robust” models, each of which makes different assumptions about the underlying nature of horizontal pleiotropy: MR-Egger regression, weighted median estimation, and weighted mode estimation^51–53^. To account for multiple testing across drug-target MR analyses, a Bonferroni correction was used (*P* < 6.17 x 10^-4^) (false positive rate = 0.05/81 statistical tests).

### Association of pre-diagnostic ANGPTL4 concentrations and colorectal cancer risk

In EPIC, blood samples were collected at recruitment and underwent proteomic analysis by Somalogic using the SomaScan 7k Assay according to the manufacturers protocol. Additional information on data pre-processing and quality control measures are presented in **Supplementary Materials**.

In analysis of directly measured pre-diagnostic ANGPTL4 concentrations, we employed Cox proportional hazard models with age as the time scale and considered “minimally adjusted” and “fully adjusted” models. Prentice weights and robust variance were used to account for the case-cohort design. Minimally adjusted models were stratified on sex, centre of origin, and age at recruitment (5-year categories). Fully adjusted models were further adjusted for body mass index (BMI), alcohol consumption, smoking, physical activity, and education level. Further information on covariate classification is presented in **Supplementary Materials**. We repeated analyses stratified by sub-site (colon cancer, rectal cancer) and tested for heterogeneity by sub-site. To explore if findings were influenced by reverse causation, we performed lag-analyses by repeating analyses excluding participants within the first 2 and 5 years of follow-up.

### Impact of ANGPTL4 loss-of-function on colon differential gene expression and gene set enrichment

To provide mechanistic insight into the effect of ANGPTL4 on pre-cancerous molecular changes within the colon, we performed a phenome-wide association study of ANGPTL4 loss-of-function on differential gene expression in normal (i.e. non-neoplastic) colon tissue samples. For these analyses, we evaluated the effect of p.E40K (rs116843064), a variant that has been shown to abolish ANGPTL4 function^54^. This variant was also employed as a genetic instrument for circulating ANGPTL4 concentrations in drug-target MR analysis.

Gene expression analysis was performed using colon biopsy RNA-seq data from the BarcUVa-Seq project^55^. This analysis was restricted to 445 individuals (mean age 60 years, 64% female, 95% of European ancestry) who participated in a Spanish colorectal cancer risk screening program that obtained a normal colonoscopy result (i.e., macroscopically normal colon tissue, with no malignant lesions). Further information on RNA-Seq and genotype data processing and quality control is presented in the **Supplementary Materials**.

Gene expression counts were normalised to account for library size differences using the trimmed mean of M-values (TMM) method^56^. Expression levels were inverse rank normal transformed. The models were adjusted for age, sex, sequencing batch, tissue location, the first two principal components of genetic ancestry, and 10 PEER factors^57^. eQTL identification was performed using linear models computed with FastQTL v211^58^. Effect estimates refer to the impact of the p.E40K minor allele (A) that causes genetic loss of ANGPTL4 function. A Benjamini-Hochberg FDR correction was used to account for multiple testing.

We then performed gene set enrichment analysis (GSEA) on genes whose expression was associated with p.E40K *(P* < 0.05) to identify biological pathways enriched among these genes using the Human MSigDB Collections Hallmark gene set^59^. The Hallmark gene set are 50 coherently expressed molecular signatures that represent well-defined biological states or processes. Gene set enrichment was performed using the fgsea R package with 1000 permutations^60^. The normalised enrichment score (NES), representing the relative enrichment of each gene set accounting for the size of the set, was calculated using the signed T-statistic as the ranking metric.

### ANGPTL4 tumour expression and all-cause mortality in colon cancer patients

To explore if ANGPTL4 is involved in cancer prognosis, we evaluated the association of *ANGPTL4* tumour expression with all-cause mortality in 481 colon cancer patients in TCGA. Read counts were normalised using the TMM method and then transformed to log2-counts per million reads. Cox proportional hazards models were employed with adjustment for age at diagnosis, gender, race, and American Joint Committee on Cancer (AJCC) pathologic stage. The time-to-event period was defined as the number of days between the initial diagnosis date and death or last follow-up. After excluding 16 participants with missing covariate data, there were 465 colon cancer patients. We did not explore the association of ANGPTL4 expression with rectal cancer because of the limited number of events (N=23) in this analysis.

This study is reported as per the STROBE-MR and STROBE reporting guidelines^61^. All statistical analyses were performed using R version 4.3.1.

## Results

### Genetic instrument validation analyses

Across the 5 drug targets, F-statistics for their instruments ranged from 306-3,388, suggesting that genetic instruments were unlikely to suffer from weak instrument bias. Characteristics of genetic variants used to proxy drug targets are presented in **Table 1**. Estimates of r^2^ and F-statistics for each target are presented in **Supplementary Table 1**.

**Table 1.** Characteristics of genetic variants used to proxy lipid-perturbing drug targets.

| Target | SNP | Effect Allele/Non-Effect Allele | Effect Allele Frequency | Beta (SE) | P-value |
| --- | --- | --- | --- | --- | --- |
| *ANGPTL3* |  |  |  |  |  |
| | rs10889352 | C/T | 0.35 | -0.27(0.01) | $< 5 \times 10^{-324}$ |
| *ANGPTL4* |  |  |  |  |  |
| | rs116843064 | A/G | 0.02 | -0.35 (0.02) | $5.70 \times 10^{-54}$ |
| *APOC3* |  |  |  |  |  |
| | rs964184 | C/G | 0.88 | -0.21 (0.01) | $2.69 \times 10^{-65}$ |
| | rs187929675 | T/C | 0.01 | -0.44 (0.04) | $2.78 \times 10^{-29}$ |
| | rs141469619 | A/G | 0.99 | -0.26 (0.04) | $4.98 \times 10^{-13}$ |
| *CETP* |  |  |  |  |  |
| | rs183130 | T/C | 0.33 | -0.32 (0.01) | $5.83 \times 10^{-136}$ |
| | rs158482 | G/T | 0.98 | -0.31 (0.05) | $1.81 \times 10^{-9}$ |
| *PCSK9* |  |  |  |  |  |
| | rs11591147 | T/G | 0.02 | -1.12 (0.02) | $< 5 \times 10^{-324}$ |
| | rs472495 | G/T | 0.36 | -0.15 (0.01) | $5.72 \times 10^{-131}$ |

Beta (SE) represents the change in circulating concentrations of the respective drug target per additional copy of the effect allele. ANGPTL3, Angiopoietin-like 3; ANGPTL4, Angiopoietin-like 4; APOC3, Apolipoprotein C3; CETP, Cholesteryl ester transfer protein; PCSK9, Proprotein convertase subtilisin/kexin type 9. Estimates were obtained from the UK Biobank for ANGPTL3, ANGPTL4, and PCSK9.

Findings from genetic instrument validation analyses were consistent with the effects of approved and emerging medications on circulating lipid biomarkers reported in clinical trials. Genetically-proxied lower ANGPTL3 (−3.10 mg/dL per SD decrease, 95% CI −3.34 to −2.86, *P* = 5.16 x 10^-146^), ANGPTL4 (−1.35 mg/dL, 95% CI −1.57 to −1.13, *P* = 6.60 x 10^-35^), and APOC3 concentrations (−0.91 mg/dL, 95% CI −1.46 to −0.36, *P* = 1.16 x 10^-3^) were associated with lower log-transformed triglyceride concentrations. Genetically-proxied lower CETP concentrations were associated with higher HDL cholesterol concentrations (1.41 µg/mL per SD decrease, 95% CI 1.14 to 1.68, *P* = 6.68 x 10^-24^). Genetically-proxied lower PCSK9 concentrations was associated with lower LDL cholesterol concentrations (−2.24 mg/dL per SD decrease, 95% CI −3.08 to −1.40, *P* = 1.37 x 10^-12^). Findings from validation MR analyses were supported in colocalisation analyses for all targets (PP_colocalisation_ > 0.80, **Supplementary Table 2**).

### Genetically-proxied drug target perturbation and cancer risk

In analysis of 78,473 cases and 107,143 controls, there was evidence that genetically-proxied circulating ANGPTL4 inhibition was associated with a reduced risk of colorectal cancer (OR per SD decrease: 0.76, 95% CI 0.66-0.89, *P* = 5.52 x 10^-4^)(**Supplementary Table 3**). There was a high posterior probability that circulating ANGPTL4 concentrations and colorectal cancer risk shared a causal variant within the *ANGPTL4* locus (PP_colocalisation_ = 0.83). In analyses stratified on colorectal cancer subsite, associations were similar across risk of colon cancer (OR 0.86, 95% CI 0.69-1.08, *P* = 0.19) and rectal cancer (OR 0.64, 95% CI 0.47-0.86, *P* = 3.20 x 10^-3^)(*P*_het_ = 0.12).

ANGPTL4 is a key regulator of plasma triglyceride levels and therefore we examined whether the association of genetically-proxied ANGPTL4 inhibition was driven by reductions in circulating triglycerides. In MR analysis, we found little evidence of association of genetically-proxied triglyceride concentrations with colorectal cancer risk in a primary IVW model (OR per unit decrease in log-transformed triglycerides: 1.04, 95% CI 0.98-1.10; *P* = 0.22) or in pleiotropy-robust models (**Supplementary Table 4**).

Genetically-proxied ANGPTL4 inhibition was not associated with risk of 5 other cancers examined (FDR *P* < 0.05). Likewise, there was no evidence of association of genetically-proxied ANGPTL3, APOC3, CETP, or PCSK9 inhibition with site-specific cancer risk (FDR *P* < 0.05)(**Supplementary Tables 5-8**). As such, subsequent analyses were restricted to ANGPTL4 and colorectal cancer and its subsites only.

### Association of pre-diagnostic ANGPTL4 concentrations and colorectal cancer risk

Case-cohort analyses included 977 incident colorectal cancer cases and 4,080 sub-cohort members (median 15.5 year follow-up) Compared to those in the lowest quartile, participants in the highest quartile of baseline circulating ANGPTL4 concentrations had higher levels of alcohol intake and were more likely to be a current smoker and to be physically active (**Table 2**). In the fully-adjusted multivariable regression model, we found evidence of a protective association of lower circulating ANGPTL4 concentrations with colorectal cancer risk (HR per log10 decrease: 0.92, 95% CI 0.85-0.99, *P* = 0.02), consistent with the genetic analyses. These findings did not differ by colorectal cancer sub-site (HR colon cancer: 0.88, 95% CI 0.81-0.96; HR rectal cancer: 0.96, 95% CI 0.85-1.08; *P*_het_ = 0.24) and were consistent in lag analyses excluding participants within the first 2 and 5 years of follow-up (**Figure 2**).

**Table 2.** Characteristics of EPIC case-cohort study participants by quartiles of circulating ANGPTL4 concentrations (N=5,057).

| Characteristic | ANGPTL4 concentrations |  |  |  |
| --- | --- | --- | --- | --- |
|  | Q1<br>(N=1265) | Q2<br>(N=1264) | Q3<br>(N=1264) | Q4<br>(N=1264) |
| Age at recruitment, y | 52.1 (9.1) | 52.8 (8.6) | 52.7 (8.7) | 52.4 (8.7) |
| Female (%) | 875 (69.2) | 790 (62.5) | 742 (58.7) | 639 (50.6) |
| Body mass index, kg/m<sup>2</sup> | 26.7 (3.9) | 27 (4.3) | 26.9 (4.3) | 27.2 (4.8) |
| Alcohol, g/day | 10.8 (16.3) | 13.1 (20.1) | 13.9 (20.4) | 16.5 (23.1) |
| Smoking (%) |  |  |  |  |
| Never | 687 (54.3) | 626 (49.5) | 600 (47.4) | 604 (47.8) |
| Former | 312 (24.7) | 321 (25.4) | 341 (27.0) | 319 (25.2) |
| Current | 266 (21.0) | 317 (25.1) | 324 (25.6) | 341 (27.0) |
| Physical activity (%) |  |  |  |  |
| Inactive | 406 (32.1) | 370 (29.2) | 368 (29.1) | 329 (26.0) |
| Moderately inactive | 402 (31.8) | 425 (33.6) | 397 (31.4) | 433 (34.3) |
| Moderately active | 220 (17.4) | 237 (18.8) | 250 (19.8) | 257 (20.3) |
| Active | 219 (17.3) | 217 (17.2) | 233 (18.4) | 243 (19.2) |
| Missing | 18 (1.4) | 15 (1.2) | 16 (1.3) | 3 (0.2) |
| Education level (%) |  |  |  |  |
| None | 236 (18.6) | 199 (15.8) | 191 (15.1) | 174 (13.8) |
| Primary | 480 (37.9) | 492 (38.9) | 452 (35.7) | 456 (36.1) |
| Secondary | 196 (15.5) | 181 (14.3) | 192 (15.2) | 172 (13.6) |
| Technical/professional | 169 (13.4) | 212 (16.8) | 197 (15.6) | 219 (17.3) |
| Longer education | 173 (13.7) | 161 (12.7) | 192 (15.2) | 187 (14.8) |
| Not specified | 11 (0.9) | 19 (1.5) | 40 (3.2) | 55 (4.4) |

Values are means and standard deviations for continuous variables and frequencies and percentages for categorical variables.

### Impact of ANGPTL4 loss-of-function on colon differential gene expression and gene set enrichment

In gene-level analysis, we did not find evidence for an association of the loss-of-function p.E40K variant with differential gene expression after correcting for multiple testing (FDR *P* < 0.05)(**Supplementary Table 9**). However, when exploring pathway-level enrichment using gene set enrichment analysis, differentially expressed genes (*P* < 0.05) were strongly enriched for 6 Hallmark gene sets (FDR *P* < 0.05). Down-regulated gene sets included those implicated in cellular proliferation (i.e. targets of the E2F family of transcription factors, genes involved in the cell cycle G2/M checkpoint, and genes involved in mitotic spindle assembly), epithelial-mesenchymal transition, and bile acid metabolism (**Figure 3**, **Supplementary Table 10**). There was one up-regulated gene set implicated in cellular proliferation (i.e. genes regulated by the oncogenic MYC pathway).

### Colon tumour ANGPTL4 expression and all-cause mortality

After a median follow-up of 1.8 (IQR 1.0-3.0) years of 465 colon cancer patients, 98 deaths were recorded. In multivariable-adjusted Cox proportional hazards models, lower colon tumour *ANGPTL4* gene expression was associated with reduced risk of all-cause mortality (HR per log2 decrease: 0.85, 95% CI 0.73-0.99; *P* = 0.04).

## Discussion

Through triangulation of evidence across proteogenomic, observational, and molecular epidemiological analyses, we prioritise ANGPTL4 as a potential therapeutic target for colorectal cancer prevention. In combined drug-target Mendelian randomization and colocalisation analyses of 78,473 cases and 107,143 controls, genetically-proxied ANGPTL4 inhibition was associated with a reduced risk of colorectal cancer. In replication analyses using an independent case-cohort of 977 incident colorectal cancer cases and 4,080 non-cases, directly measured lower circulating ANGPTL4 concentrations were associated with reduced colorectal cancer risk. In gene set enrichment analysis of differential gene expression in 445 normal colon tissue samples, ANGPTL4 loss-of-function was associated with down-regulation of several cancer-related gene-sets. Finally, in analysis of 465 colon cancer patients, lower *ANGPTL4* expression in colon tumour tissue was associated with a reduced risk of all-cause mortality. Collectively, these findings provide strong and consistent support for a role of ANGPTL4 in colorectal tumorigenesis.

ANGPTL4 is a ubiquitously expressed glycoprotein that inhibits lipoprotein lipase and modulates fatty acid uptake in adipose and oxidative tissue^62–66^. As a key regulator of triglyceride clearance, ANGPTL4 has emerged as an attractive therapeutic target for reducing triglyceride levels and adverse cardiovascular events^67,68^. This is supported by human genetic evidence that loss-of-function variants in *ANGPTL4* are associated with lower plasma triglycerides and reduced coronary artery disease risk^69^. Human genetic inactivation of *ANGPTL4* has also been shown to improve glucose homeostasis and reduce type 2 diabetes risk and *Angptl4* deletion in mice has been reported to improve insulin sensitivity and glucose homeostasis in fasting and postprandial states^70^. At least two pharmacological ANGPTL4 inhibitors are currently under Phase II clinical trial evaluation for their efficacy in lowering plasma triglycerides and reducing cardiovascular events^71,72^.

Our findings implicating ANGPTL4 in colorectal cancer development recapitulate insights from preclinical studies. For example, *ANGPTL4* knockdown has been shown to inhibit proliferation, promote apoptosis, and suppress migration in colorectal cancer cell lines and to reduce colorectal tumour size in xenograft mouse models^73,74^. Recombinant ANGPTL4 has been reported to promote colon cancer growth by impairing CD8+ T cell activity in mice^75^. Recently, ANGPTL4 suppression has been shown to reprogram endothelial cell metabolism and inhibit angiogenesis, providing another mechanism through which ANGPTL4 may influence carcinogenesis^76^. Interestingly, prostaglandin E2, a putative key mediator of the effect of COX-2 on colorectal cancer, has also been reported to promote colorectal carcinoma cell proliferation via ANGPTL4 under hypoxic conditions^77,78^. Consistent with prior reports, we did not find evidence of an association of genetically-proxied triglyceride concentrations with colorectal cancer risk, suggesting that the association between ANGPTL4 and colorectal cancer risk is mediated via pathways independent of triglyceride lowering^26,79^. In gene set enrichment analysis, ANGPTL4 loss-of-function lead to down-regulation of several biological pathways implicated in colon carcinogenesis including cellular proliferation, bile acid metabolism, and the epithelial-mesenchymal transition. For example, bile acids have been shown to promote colon cancer by damaging colonic epithelial cells, and inducing reactive oxygen species production, genomic destabilisation, and apoptosis resistance^80^. In addition, the epithelial-mesenchymal transition (EMT) has been reported to play an important role in colorectal cancer progression, metastasis, and drug resistance, and preclinical studies have suggested the efficacy of pharmacological perturbation of markers of the EMT in colorectal cancer^81^. Our findings thus validate and extend insights from preclinical cancer models and can help to guide future work investigating mechanisms underpinning the effect of ANGPTL4 on colorectal cancer development.

Contrary to some prior studies, we found little evidence to support associations of other lipid-perturbing targets with cancer risk. For example, we failed to detect previously reported associations between genetically-proxied PCSK9 inhibition and site-specific cancer risk (i.e. breast, prostate, head and neck)^27,28,82^. In addition, we did not find evidence to support previously reported adverse effects of genetically-proxied CETP inhibition on breast cancer risk^82^. The absence of or inconsistent application of colocalisation analysis in some prior analyses complicates assessment of whether discordance between findings reflects the presence of confounding by LD in previous studies, differences in instrument construction strategy across studies, or chance. Nonetheless, our findings suggesting little evidence of association of genetically-proxied ANGPTL3, APOC3, CETP, and PCSK9 inhibition with cancer risk may help to deprioritise further evaluation of these proteins as intervention targets for cancer prevention.

Strengths of this study include use of a triangulation framework leveraging genetic and conventional epidemiological approaches to strengthen causal inference. Notably, the consistency of findings across drug-target Mendelian randomization and conventional epidemiological analysis, both of which may be susceptible to unrelated sources of bias, permitted us to increase confidence in our conclusions relating circulating ANGPTL4 to colorectal cancer risk^83^. By leveraging gene expression data from normal and cancerous colon tissue samples we were able to gain mechanistic insight into the effect of ANGPTL4 on early precancerous changes in the colon and to extend exploration of the role of ANGPTL4 to mortality among colon cancer patients, supporting a role of this target across the carcinogenesis spectrum.

There are several limitations to this analysis. First, drug-target MR analyses assume exchangeability and exclusion restriction. While various sensitivity analyses were performed to evaluate the robustness of findings to violations of both assumptions, these are unverifiable.

Second, drug-target MR estimates assume linear and time-fixed effects and the absence of gene-gene or gene-environment interactions. Third, conventional observational analyses performed in EPIC and TCGA assume the absence of confounding, measurement error, and reverse causation though lag analyses in EPIC were consistent with the primary analysis. Fourth, genetic and conventional observational analyses are restricted to examining on-target (i.e. target-mediated) effects of medications. Fifth, statistical power was likely limited in drug-target MR analyses of less common cancer subtypes. Sixth, genetic and conventional observational analyses were primarily performed in participants of European ancestry and, therefore, the generalisability of these findings to non-European populations is unclear. Seventh, we were unable to explore the association of both *ANGPTL4* loss-of-function with differential gene expression in normal rectal tissue because of the absence of suitable data in this tissue and *ANGPTL4* expression in rectal tumour samples with all-cause mortality due to the limited number of events in this dataset.

Colorectal cancer is the third most common cancer globally, accounting for over 900,000 deaths in 2022^84,85^. Aspirin and nonsteroidal anti-inflammatory drugs can be used to lower colorectal cancer risk in high-risk populations (e.g. individuals with Lynch syndrome, familial adenomatous polyposis) but the increased risk of gastrointestinal bleeding on these medications limit their wider use^86^. There is therefore a need for identification of novel safe and effective chemoprevention agents for colorectal cancer to reduce the burden from this disease. Our findings leveraging genetic, observational, and molecular epidemiological designs recapitulate insights from preclinical studies indicating a protective effect of ANGPTL4 inhibition in colorectal cancer risk. Further work validating findings in human studies and clarifying potential mechanisms of effect will guide further assessment of the viability of ANGPTL4 inhibition as a therapeutic strategy for cancer prevention. In addition, investigation of the role of ANGPTL4 in colorectal carcinogenesis in non-European populations will permit evaluation of the generalisability of these findings to other ancestries. Finally, ongoing clinical trials investigating pharmacological ANGPTL4 inhibition for CVD present another opportunity to explore potential cancer preventive properties of these medications.

## Conclusion

Our comprehensive proteogenomic and observational analyses suggest a protective role of lowering circulating ANGPTL4 concentrations in colorectal cancer risk. These findings provide human validation to insights from preclinical studies and support the further evaluation of ANGPTL4 as a potential therapeutic target for colorectal cancer prevention.

## Figure footnotes

**Figure 2**

2-year lag = removed participants within the first 2 years of follow-up, 5-year lag = removed participants within the first 5 years of follow-up

Minimally adjusted model was stratified on sex, centre of origin, and age at recruitment. The fully adjusted model was further adjusted for body mass index (BMI), alcohol (grams/day), smoking status (current, former, never smoker, unknown), physical activity index (inactive, moderately inactive, moderately active, active, missing), and highest level of education (not specified, none, primary, secondary, technical/professional, longer education).

## Funding

JY and IT are supported by the National Institute for Health and Care Research Imperial Biomedical Research Centre. DW and EL are supported by A*STAR (UIBR), the Academy of Medical Sciences Professorship (APR7_1002) and the Engineering and Physical Sciences Research Council (EP/V029045/1). FMN and VM are supported by the Spanish Association Against Cancer (AECC) Scientific Foundation grant GCTRA18022MORE, the Consortium for Biomedical Research in Epidemiology and Public Health (CIBERESP), action Genrisk and the Instituto de Salud Carlos III (ISCIII), “Programa FORTALECE del Ministerio de Ciencia e Innovación”, through the project number FORT23/00032.

## Data Availability

All data produced are available online except for case-cohort analyses performed in the European Prospective Investigation into Cancer and Nutrition (EPIC) which required submission of a data usage proposal.

## Acknowledgements

The authors thank the National Institute for Public Health and the Environment (RIVM), Bilthoven, the Netherlands, for their contribution and ongoing support to the EPIC Study. The authors also thank CERCA Programme, Generalitat de Catalunya for institutional support.

## Disclaimer

Where authors are identified as personnel of the International Agency for Research on Cancer/World Health Organization, the authors alone are responsible for the views expressed in this article and they do not necessarily represent the decisions, policy or views of the International Agency for Research on Cancer/World Health Organization.

---

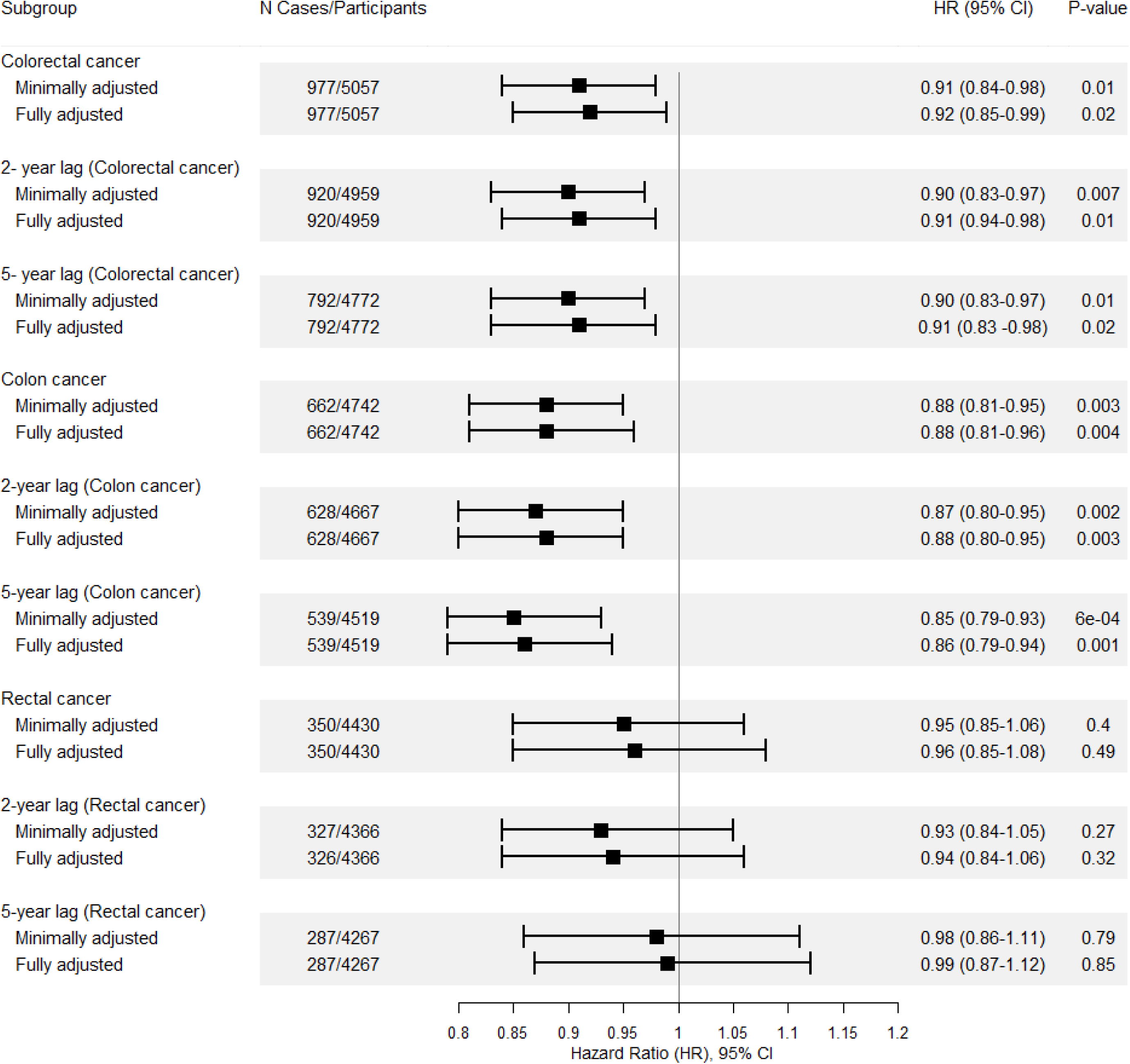

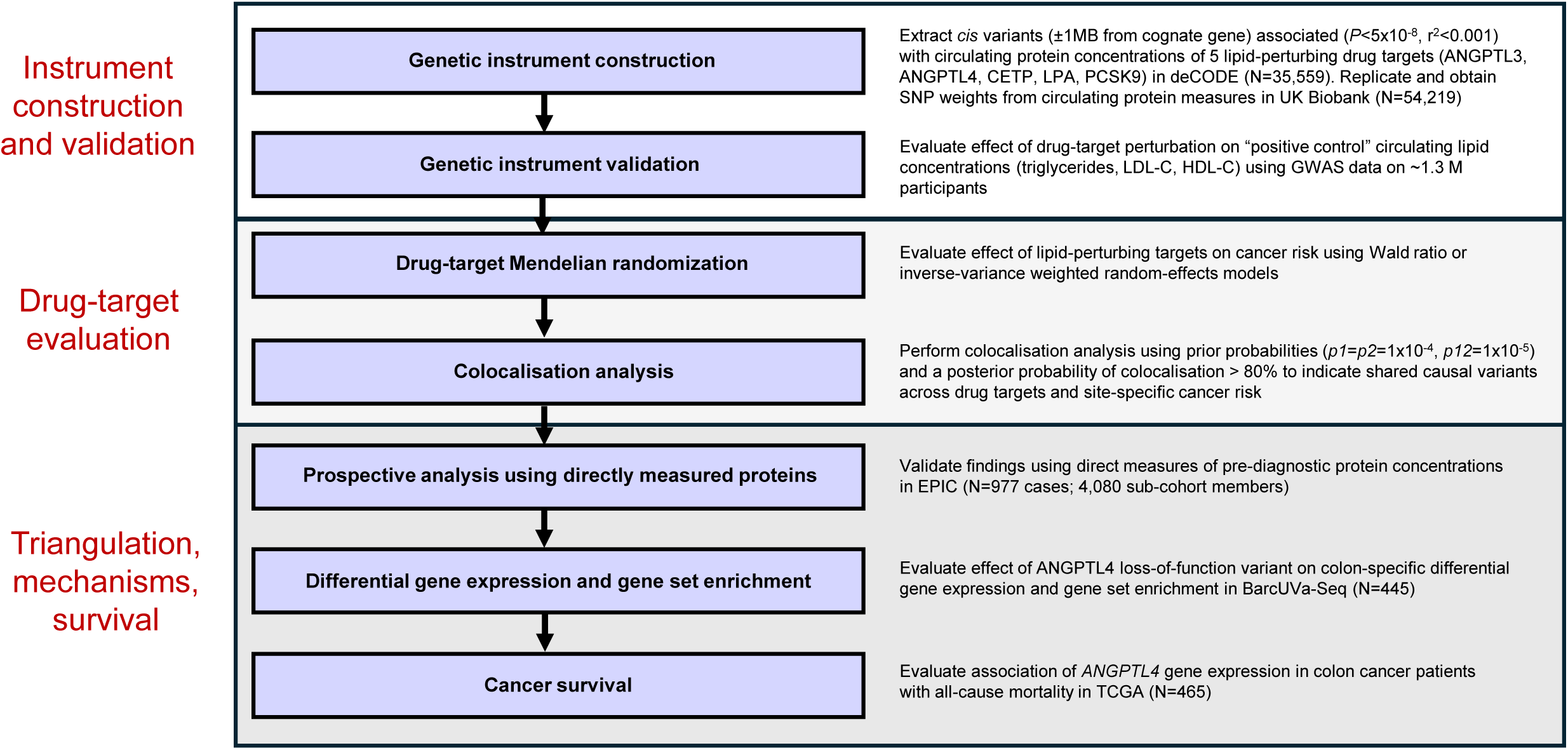

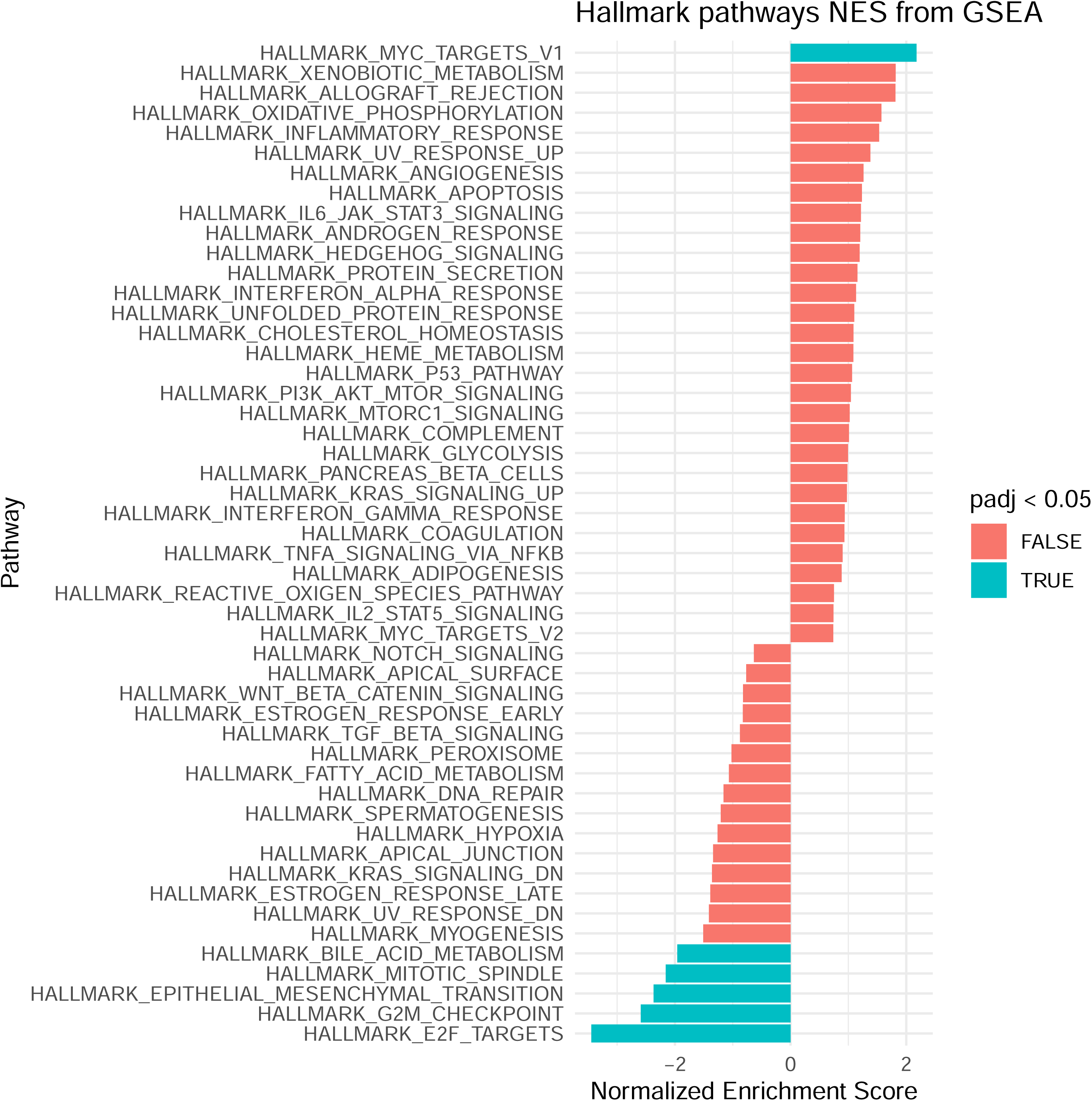

## Notes

### Competing Interest Statement

The authors have declared no competing interest.

### Author Declarations

Access to European Prospective Investigation into Cancer and Nutrition (EPIC) data was granted following submission of a data usage proposal.

