## Supplementary Materials for "Proteogenomic and observational evidence implicate ANGPTL4 as a potential therapeutic target for colorectal cancer prevention"

**Validation analysis using direct protein measures**

Assessment of exposure

Blood samples were collected at recruitment and underwent proteomic analysis by Somalogic using the SomaScan 7k Assay according to the manufacturers protocol. Briefly, the SomaScan platform uses modified nucleotides (Slow Off-rate Modified Aptamers; SOMAmer) which make direct contact with proteins, enabling detection of proteins or protein complexes and quantifies them in relative fluorescence units (RFUs) using DNA microarray^2^. Separate SOMAmers can bind to isoforms of the same protein but can also bind to the same protein at different sites (which can be impacted by post-translational modifications or complexes formed with other proteins). RFUs were first normalized by the manufacturer through the following steps: hybridization normalization, intraplate median normalization, plate scaling and calibration, and adaptive normalization to a population reference. We: i) log-transformed these normalized RFUs to reduce skewness; ii) excluded samples where the normalization scale factor was outside [0.4-2.5]; iii) excluded samples using principal component analyses (PCA) and a local outlier factor using a Tukey rule modified to account for skewness and multiple testing^1^; iv) applied plate correction using a residual approach whereby, for each SOMAmer, its measurements were corrected for plate effect estimated in linear mixed effect models adjusted for centre, age, sex, BMI, smoking status, and incidence of cancer, CVD, T2D and death, to preserve possible biological variation due to these factors^2^; v) centred and scaled measurements of each SOMAmer so that their mean and standard deviation in the sub-cohort were 0 and 1.

Multi-endpoint case-cohort design in EPIC

A multi-endpoint case-cohort was established within EPIC in which individuals recruited in the UK, Netherlands, Spain, and Italy were eligible. Individuals were excluded if they had prevalent cancer or cardiovascular disease, had blood collection <35 or >75 years, or missing data for lifestyle or reproductive factors. Individuals were then included if they: were included in the EPIC-InterAct subcohort^3^, had a major cancer, or were a randomly selected type 2 diabetes, coronary heart disease, stroke, or death case. In the present study, we focus on individuals included in the EPIC-InterAct sub-cohort and/or the cancer case component which included a total of 10,261 individuals.

Covariate classification

Age at event was defined as age at diagnosis or end of follow-up; 5-year age groups were defined from 0 to the maximum age of the included participants; BMI was measured as weight (kg) divided by height (m^2^); alcohol consumption was self-reported as grams/day; smoking was self-reported as any one of: never, former, current, unknown (never was used as the reference); physical activity was self-reported using the Cambridge physical activity index as any one of: inactive, moderately inactive, moderately active, active, missing (inactive was used as the reference; categorical); education level was self-reported as any one of: none, primary school completed, secondary school, technical/professional school, longer education (incl. University degree), not-specified (none was used as the reference).

**Impact of ANGPTL4 loss-of-function on colon differential gene expression and gene set enrichment**

RNA-Seq processing

The raw RNA-Seq reads were processed using the BBTools suite to remove low-quality bases, adapters, and residual ribosomal RNA sequences^4^. The trimmed reads were then aligned to the human reference genome GRCh37 using STAR v2.5, with annotations from GENCODE release 19^5,6^. For further analysis, only samples with more than 10 million mapped paired-end reads, a unique mapping rate greater than 80%, and a multimapping rate lower than 15% were included. Gene expression was quantified using RSEM^7^.

Genotype processing

Blood samples were genotyped using the Illumina OncoArray BeadChip^8^. The quality control criteria included the following: i) genotyping rate > 95%, ii) relatedness > 0.8, iii) SNP and per-sample missing rate > 0.1, iv) sex concordance between genetic data and reported information. Imputation was performed using the Haplotype Reference Consortium panel via the Michigan Imputation Server^9^. 6.9 million SNPs were retained after filtering for a minor allele frequency lower than 1% and an R 2 quality score greater than 0.7. SNP identifiers were annotated with dbSNP v142^10^. The concordance between genotype and RNA-seq samples was verified using CheckFingerprint^11^.

**Supplementary Materials Citations**

1	Breunig, M. M., Kriegel, H.-P., Ng, R. T. & Sander, J. in *Proceedings of the 2000 ACM SIGMOD international conference on Management of data*	93–104 (Association for Computing Machinery, Dallas, Texas, USA, 2000).

2	Viallon, V. *et al.* A New Pipeline for the Normalization and Pooling of Metabolomics Data. *Metabolites* **11** (2021). <https://doi.org/10.3390/metabo11090631>

3	Forouhi, N. G. & Wareham, N. J. The EPIC-InterAct Study: A Study of the Interplay between Genetic and Lifestyle Behavioral Factors on the Risk of Type 2 Diabetes in European Populations. *Curr Nutr Rep* **3**, 355-363 (2014). <https://doi.org/10.1007/s13668-014-0098-y>

4	*BBMap. SourceForge.*, <<https://sourceforge.net/projects/bbmap/>> (

5	Dobin, A. *et al.* STAR: ultrafast universal RNA-seq aligner. *Bioinformatics* **29**, 15-21 (2013). <https://doi.org/10.1093/bioinformatics/bts635>

6	Harrow, J. *et al.* GENCODE: the reference human genome annotation for The ENCODE Project. *Genome Res* **22**, 1760-1774 (2012). <https://doi.org/10.1101/gr.135350.111>

7	Li, B. & Dewey, C. N. RSEM: accurate transcript quantification from RNA-Seq data with or without a reference genome. *BMC Bioinformatics* **12**, 323 (2011). <https://doi.org/10.1186/1471-2105-12-323>

8	Amos, C. I. *et al.* The OncoArray Consortium: A Network for Understanding the Genetic Architecture of Common Cancers. *Cancer Epidemiol Biomarkers Prev* **26**, 126-135 (2017). <https://doi.org/10.1158/1055-9965.Epi-16-0106>

9	Das, S. *et al.* Next-generation genotype imputation service and methods. *Nat Genet* **48**, 1284-1287 (2016). <https://doi.org/10.1038/ng.3656>

10	Database resources of the National Center for Biotechnology Information. *Nucleic Acids Res* **44**, D7-19 (2016). <https://doi.org/10.1093/nar/gkv1290>

11	*picard/src/main/java/picard/fingerprint/CheckFingerprint.java at master · broadinstitute/picard. GitHub*, <<https://github.com/broadinstitute/picard/blob/master/src/main/java/picard/fingerprint/CheckFingerprint.java>> (
